# Can synthetic data overcome the privacy and fidelity bottleneck in Pharmacometrics? A comparative benchmark using a daptomycin population pharmacokinetic model

**DOI:** 10.64898/2026.05.30.26354512

**Authors:** Alexandre Destere, Romain Lombardi, Marc Labriffe, Clément Benoist, Pierre Marquet, Thibaud Lavrut, Alexandre Gérard, Charles Bouveyron, Jean-Baptiste Woillard

## Abstract

**Introduction:** The sharing of individual patient data is essential for advancing pharmacometrics but is strictly limited by privacy regulations (e.g., GDPR). While synthetic data generation offers a legally compliant alternative, its structural impact on complex nonlinear mixed-effects (NLME) modelling remains largely unexplored. This study aimed to benchmark five generative artificial intelligence algorithms by evaluating the balance between data privacy and the preservation of structural PK properties and clinical dosing guidance.

**Material & methods:** A daptomycin two-compartment PopPK model was used to simulate a reference cohort of 500 patients. Five generative algorithms (Modified AVATAR, Gaussian Copula, Synthpop, TVAE, and CTGAN) produced 100 independent synthetic datasets each. A two-stage evaluation framework was applied: first, a statistical indistinguishability test based on logistic regression (AUC ROC) was used as a macroscopic pre-selection criterion to determine algorithm eligibility for NLME modelling and privacy risk assessment. Privacy risk was independently quantified using the Anonymeter framework (Singling Out and Linkability attacks). Eligible algorithms were further evaluated on PK parameter recovery bias and clinical dosing simulations.

**Results:** Deep learning architectures (TVAE, CTGAN) were excluded at the pre-selection stage due to both biologically implausible covariate generation and high macroscopic detectability (mean AUC ROC = 0.837 and 0.986, respectively). Synthpop, AVATAR, and Gaussian Copula all passed the indistinguishability threshold (AUC ROC = 0.475 ± 0.033, 0.490 ± 0.013, and 0.619 ± 0.031, respectively) and proceeded to NLME evaluation. However, attack-based privacy assessment revealed that Synthpop carried an unacceptable singling-out risk (0.035), disqualifying it from privacy-preserving data sharing. AVATAR and Gaussian Copula demonstrated acceptable privacy profiles (singling-out = 0.004 and 0.001; linkability = 0.010 and 0.003, respectively). At the structural level, Gaussian Copula injected stochastic noise inflating residual error (+157.0%) and VLJ (+25.9%), blunting predicted Cmax and predisposing to empirical dose escalation and risk of toxicity. AVATAR acted aSs a smoothing filter, deflating VLJ (−48.3%) and underestimating CL (−12.9%). Forward clinical simulations confirmed directionally opposed prediction errors: Gaussian Copula consistently underestimated Cmax across standard and renally impaired profiles (−14.5% and -16.0%, respectively), predisposing to empirical dose escalation, whereas AVATAR- and Synthpop-derived models overestimated Cmax and Cmin in the obese infected patient (+14.7% and +8.2%, respectively), compounding the accumulation risk already present in this profile.

**Conclusion:** While no generative algorithm currently offers a perfect solution, AVATAR and Gaussian Copula represent the most viable candidates, being the only methods to satisfy both macroscopic indistinguishability and attack-based privacy criteria. These findings highlight the necessity of a structured, two-stage validation framework and suggest that, when coupled with therapeutic drug monitoring, synthetic datasets could significantly enhance multicentre collaboration while maintaining strict regulatory compliance.

## Introduction

The production and acquisition of clinical data in pharmacology are complex processes, often impeded by confidentiality issues, particularly in Europe where the General Data Protection Regulation (GDPR) imposes strict restrictions. An Australian study highlighted these vulnerabilities, demonstrating that up to 10% of the data contained in open-access databases could be used to re-identify patients [1]. In light of these challenges, one potential solution rapidly gaining traction is the creation of synthetic data. Designed to closely mimic the multidimensional characteristics of real clinical data, synthetic data preserves the integrity and utility of the original information for research while mathematically avoiding the possibility of patient re-identification [2].

The sharing of such synthetic data between partner centres involved in multicentric projects could drastically facilitate collaboration, a crucial step for advancing Population Pharmacokinetics (PopPK) and optimizing Model-Informed Precision Dosing (MIPD). To achieve this, a variety of generative techniques have been employed, ranging from statistical modelling to deep learning. Among these, the “AVATAR” method involves dimensionality reduction through the utilization of the k-nearest neighbours (KNN) method in a latent space, establishing a localized modelling area for each patient [3]. Other state-of-the-art methods include Tabular Variational Autoencoders (TVAE), which use an encoder-decoder structure with Gaussian noise in the latent space, and Conditional Tabular Generative Adversarial Networks (CTGAN), which train a generator and a discriminator simultaneously. Furthermore, statistical approaches like Gaussian Copulas [4], which function by isolating marginal distributions from their underlying multivariate correlation structure and sequential synthesis methods (for instance, Synthpop [5]), which generate synthetic data variable-by-variable using conditional regression models, offer alternative frameworks for capturing complex variable distributions [6,7].

Despite their success in cross-sectional tabular data, these generative approaches are rarely used in the field of population pharmacokinetics. PopPK data presents unique challenges: it is intrinsically longitudinal, often sparse, and governed by underlying physiological mechanisms described by complex ordinary differential equations (ODEs) [8]. Many commonly used synthetic data generation methods in healthcare, particularly deep learning–based models, require large training datasets to learn the underlying data distribution reliably [9], a prerequisite often difficult to fulfil in PopPK studies. Consequently, generating synthetic PopPK datasets carries the significant risk of altering the underlying kinetic relationships, which may result in a change to the selected structural model or introduce severe biases in the estimation of typical PK parameters and inter-individual variability.

Furthermore, a major but frequently overlooked limitation of generative algorithms is their inherent stochasticity. Generating a single synthetic dataset relies heavily on the random seed used, leading to potential intra-algorithmic variability. To truly validate a generative method for pharmacometrics, its stability and reproducibility must be objectively quantified through repeated generation cycles.

Therefore, the aim was to benchmark five generative algorithms (Modified AVATAR, TVAE, CTGAN, Gaussian Copula, and Synthpop) for their ability to produce synthetic longitudinal PopPK data, using a well-established daptomycin two-compartment model as a controlled ground truth. To ensure both reproducibility and robust characterisation of intra-algorithmic variability, 100 independent synthetic datasets were generated per algorithm. Algorithm performance was evaluated through a structured, two-stage framework: first, a macroscopic statistical indistinguishability screen based on logistic regression (AUC ROC) was applied as an eligibility filter for downstream NLME inference; second, a mechanistic, attack-based privacy assessment using the Anonymeter framework (Singling Out, Linkability, and Inference) was conducted independently to quantify re-identification risk. Algorithms satisfying both criteria were then subjected to rigorous nonlinear mixed-effects re-estimation to quantify structural PK parameter bias, and to forward clinical simulations to assess the potential impact on Model-Informed Precision Dosing.

## Material and methods

All the statistical analyses and plots generations were performed either in R software 4.5.1 or in Python software 3.12.

### In Silico Framework and Reference Dataset

To provide a robust and known ground truth for evaluating synthetic data generation methods, a Stochastic Simulation and Estimation framework was employed. The reference dataset was generated via stochastic simulation using the R package *mrgsolve* (v 1.5.2) [10], based on a previously published and well-characterized multi-compartmental population pharmacokinetic (PopPK) model of daptomycin [11].The structural model was a two-compartment model with first-order elimination from the central compartment. The typical population parameters (0_TV_ ) were defined as follows: clearance (CL) = 0.807 L/h, central compartment volume (V1) = 4.80 L, inter-compartmental distribution clearance (Q) = 3.46 L/h, and peripheral compartment volume (V2) = 3.13 L. Between-subject variability (BSV) was modelled using an exponential distribution to ensure the strict positivity of individual parameters:

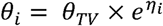

Where 0_i_ is the parameter for the i-th individual, 0_TV_ is the typical population value, and is a random variable drawn from a normal distribution with a mean of zero and variance w^2^. The estimated variances (w^2^) were 0.0905, 0.2740, 0.3561, and 0.0359 for CL, V1, Q, and V2, respectively. Unexplained residual variability (RUV) was described by a strictly additive error model (er = 2.08 mg/L).

To reproduce a realistic clinical population similar to that reported in the original article, several covariates–parameter relationships were incorporated into the underlying differential equations. CL was modelled as a function of estimated creatinine clearance (CLCR, calculated using the Cockcroft–Gault equation), body temperature (TEMP), and sex (SEX), with sex coded proportionally as 0.8 for females and 1.0 for males. No covariate effect was applied to V1. Q was modelled with a linear effect of body weight (WT) relative to the reference value of 75.1 kg, while V2 included both a linear effect of WT deviation and a multiplicative effect related to the presence of systemic infection (INF).

A simulated cohort of 500 patients based on this PopPK model were created. Continuous covariates were sampled from truncated normal distributions (WT, Age, TEMP) or log-normal distributions (serum creatinine), while categorical covariates (sex, infection status) were drawn from binomial , with prevalences as reported in the original article. Each virtual patient received a standard daptomycin dosing regimen of 10 mg/kg administered as a 30-minute intravenous infusion every 24 hours. Concentration-time profiles were simulated at steady-state, capturing data between 120 and 144 hours post-initiation.

### Synthetic Data generation algorithms

Because generative algorithms are primarily designed for cross-sectional data, the simulated longitudinal reference data were first converted into a wide format (one row per patient, with distinct columns for concentrations at each sampled time point). These formatted data were then subjected to five distinct generative frameworks. To thoroughly assess the stability, reproducibility, and intra-algorithmic variability of each method, 100 independent synthetic datasets (N=500 virtual patients per dataset) were generated for each of the five algorithms.

### Modified **“**AVATAR**”** Synthesis

A custom stochastic algorithmic method based on the methodology proposed by Guillaudeux *et al.* [3] which was recently endorsed by the French regulatory body for data privacy (CNIL), was adapted for this study. The generation process involved: (i) dimensionality reduction: Application of Principal Component Analysis (PCA) on the standardised data matrix. (ii) dynamic neighbourhood optimisation:

For each individual i, the k-nearest neighbours (KNN) algorithm was applied within the PCA latent space. The optimal number of neighbours (k) was automatically and dynamically determined by identifying the knee point on the curve of the median intra-neighbourhood Euclidean distance. (iii) stochastic generation: The synthetic twin (AVATAR) was generated through a weighted linear combination of its k neighbours’ vectors. The weights integrated the inverse of the Euclidean distance, modulated by stochastic noise drawn from an exponential distribution, and an attenuation factor linked to the neighbour’s rank (0.5^rank^). (iv) reverse engineering: The synthetic latent coordinates underwent an inverse PCA transformation, de-standardisation, and probabilistic thresholding for binary covariates (0.5 threshold), before being reformatted into the standard longitudinal PopPK format.

#### Machine Learning and statistical models

Four additional state-of-the-art generative methods were applied for benchmarking: CTGAN, TVAE[7], Gaussian Copula [4], implemented using the *SDV* Python library [12] and Synthpop, implemented via the *synthpop* R package (v 1.9-2) [5].

### Analysis of statistical indistinguishability and Algorithm pre-selection

To determine whether synthetic datasets were macroscopically representative of the original data distribution, a logistic regression classifier was trained for each generative algorithm to predict whether a patient record originated from the real or synthetic cohort. A logistic regression classifier was deliberately selected over more complex algorithms to ensure interpretability and avoid overfitting on a 500-patient dataset. In this adversarial framework, a simple linear classifier provides a conservative lower bound on dataset separability: if even a logistic model can distinguish real from synthetic records, the structural discrepancy is unambiguously macroscopic and clinically meaningful. This analysis was not designed as a privacy assessment *per se*, but rather as a global structural screening tool: an algorithm producing synthetic data that remains statistically indistinguishable from the original at the population level provides a meaningful basis for downstream NLME inference, whereas one generating easily separable data reflects macroscopic structural distortions that would inevitably propagate into and compromise pharmacokinetic parameter estimates.

A total of 100 classifiers per generative algorithm were trained. For each iteration, the original reference dataset and the corresponding synthetic dataset were combined and assigned a binary target variable (”Real” vs. “Synthetic”). The pooled data were randomly partitioned into training (75%) and test (25%) sets using stratified sampling. A standardised preprocessing pipeline was applied, including removal of zero-variance predictors, centring and scaling of continuous variables, and one-hot encoding of categorical variables. Discriminative performance was evaluated using the AUC ROC on the held-out test set. An AUC approaching 0.5 indicates near-perfect macroscopic indistinguishability; an AUC approaching 1.0 reflects structural discrepancies that would compromise downstream NLME validity. A pre-specified threshold of AUC ROC = 0.75 was used as the eligibility criterion for proceeding to PopPK modelling. Intra-algorithmic stability was characterised by the mean, standard deviation, median, and interquartile range across 100 iterations. A threshold of 0.75 was selected a priori as a clinically meaningful midpoint between chance-level performance (0.5) and near-perfect separation (>0.9), representing a level of distributional overlap deemed insufficient for valid pharmacokinetic inference.

### Population Pharmacokinetic Modelling and Inference Evaluation

The primary endpoint was the preservation of the initial pharmacokinetic conclusion. The synthetic data were converted back to the standard longitudinal PopPK format. For each algorithm, 100 synthetic datasets were generated, and for each dataset, 0_TV_ ) and the inter-individual variabilities terms of the PopPK model were re-estimated using the First-Order Conditional Estimation with Interaction (FOCEi) algorithm implemented in the *nlmixr2* R package (v 4.0) [13]. Importantly, while the structural parameter (0_TV_ ) and inter-individual variabilities were re-estimated, the covariate coefficients were fixed. This strategy was chosen to isolate the ability of the generative algorithms to preserve the underlying compartmental kinetics and baseline variance structure, while avoiding confounding from re-estimation of covariate effects, which could both compensate for poorly recovered structural parameters and introduce additional variability.

The utility of each generative method was quantified by calculating the relative mean predictive error (rMPE%) for each structural parameter (CL, V1, Q, V2) and the residual error model (er), comparing the estimates derived from the synthetic data to the reference parameters of the original simulated population:

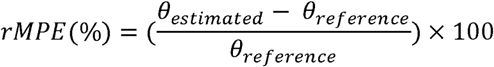

To establish a rigorous comparative baseline and isolate the bias specifically introduced by the generative algorithms, the initial simulated dataset used to generate the synthetic data was also analysed using the same population re-estimation procedure (FOCEi). This preliminary analysis made it possible to quantify the estimation bias arising from the structural model and sampling design alone, independently of any synthetic data generation process. Therefore, the performance of the synthetic datasets was evaluated by calculating the relative bias of their estimated parameters relative to these original re-estimated values, rather than solely relative to the initial simulation parameters.

To quantify intra-algorithmic variability, the standard deviations and 95% confidence intervals of the estimated parameters were calculated across the 100 iterations. Methods that failed to reach algorithmic convergence (such as specific neural network architectures that disrupted the required longitudinal ODE structure) were explicitly tracked and excluded from the mean bias calculations. Data visualization was performed using the *ggplot2* R package (v 4.0.1) to visually compare the discrepancy across the evaluated algorithms.

### Privacy Risk Assessment

To complement the adversarial AUC ROC validation with a mechanistic, attack-based quantification of re-identification risk, privacy was further assessed using the Anonymeter library (v1.0.0) [14] across three paradigms: Singling Out, Linkability, and Inference.

All analyses were performed on 100 independent subsets (dataset_indices) of 500 individuals each, drawn from the pooled synthetic populations. The original simulated cohort (ori) served as the training reference, and an independent real dataset (control) derived from a second independent simulation run was used as the holdout reference. The control dataset was pre-processed by removing uninformative columns and realigning its variable structure to match ori.

Singling Out was evaluated in multivariate mode using a “Singling Out Evaluator”(n of attacks = 500), applied across three random seeds (42, 123, 456) per subset. The privacy risk for each subset was defined as the mean across seeds, and a global estimate was obtained as the mean across the 100 subsets.

Linkability risk was assessed by evaluating whether an adversary who independently holds two partial views of the dataset — a demographic profile (AMT, WT, CREAT, AGE, SEX, TEMP, INF) and a pharmacokinetic profile (26 steady-state concentration variables, conc_120 to conc_144) — could successfully link a given individual across these two views using the synthetic data as a bridge. A single evaluation pass per subset was performed (n_attacks = 500); because the number of attacks matched the subset size, all individuals were targeted, eliminating stochasticity. Inference was evaluated using an “Inference Evaluator” applied in a double loop across subsets and columns (n of attacks = 500). For all three paradigms, 95% confidence intervals were derived by non-parametric bootstrapping (n_resamples = 9,999, random state = 42).

### Evaluation of Clinical Implications and Predictive Performance

Forward deterministic simulations were performed to translate structural parameter biases into clinically meaningful outcomes and to evaluate the a priori suitability of synthetic-data-derived models for Model-Informed Precision Dosing (MIPD). Three virtual patient profiles were considered: a standard adult (75 kg, creatinine clearance [CrCl] 91 mL/min, no infection), a frail low-weight adult (45 kg, CrCl 30 mL/min) at high risk of drug accumulation, and an obese adult (120 kg, CrCl 150 mL/min) with systemic infection at high risk of underexposure. For the original reference population and each convergent generative algorithm (AVATAR, Gaussian Copula, and Synthpop), FOCEi-estimated typical population parameters were used to simulate steady-state pharmacokinetic profiles under daptomycin 10 mg/kg every 24 hours administered as a 30-minute intravenous infusion. Simulations were conducted over 144 hours. Performance was assessed using steady-state Cmax and Cmin, which were interpreted according to established daptomycin therapeutic thresholds for efficacy and skeletal muscle toxicity (Cmin > 24.3 mg/L).

## Results

### Synthetic Data Generation and Population Characteristics

A total of 100 independent synthetic datasets of 500 patients were successfully generated for each of the five evaluated algorithms (AVATAR, Gaussian Copula, Synthpop, TVAE, and CTGAN). To evaluate the global generative tendencies and isolate extreme values, these iterations were pooled, resulting in a cumulative virtual population of 50,000 patients per method. The demographic, clinical, and raw pharmacokinetic characteristics are summarised in Table 1. Visual and statistical inspection of the synthetic populations immediately highlighted stark disparities between the generative frameworks (Figure 1). The modified AVATAR, Gaussian Copula, and Synthpop successfully preserved the macroscopic physiological structure of the original cohort. Median age, weight, and initial CrCl remained closely aligned with the original population (e.g., median CrCl: 140.1 mL/min for simulated original vs. 160.3, 130.7, and 167.7 mL/min for AVATAR, Copula, and Synthpop, respectively). Conversely, deep learning architectures (TVAE and CTGAN) demonstrated severe limitations in respecting biological and physiological boundaries within the constraints of our sample size (n = 500). TVAE significantly distorted the demographic balance, increasing the male proportion from 59% to 75% and shifting the median age to 43.2 years. More critically, CTGAN generated biologically implausible covariate distributions, notably a hyper-metabolic median CrCl of 253.4 mL/min, with the 75th percentile reaching 446.3 mL/min. Consequently, during the structural re-estimation step, all 100 datasets generated by TVAE and CTGAN systematically failed to converge. This failure was driven by extreme physiological aberrations and a lack of continuous constraints, which prevented the FOCEi algorithm from resolving the underlying ODE. They were therefore excluded from further pharmacokinetic and clinical evaluation.

**Figure 1:**
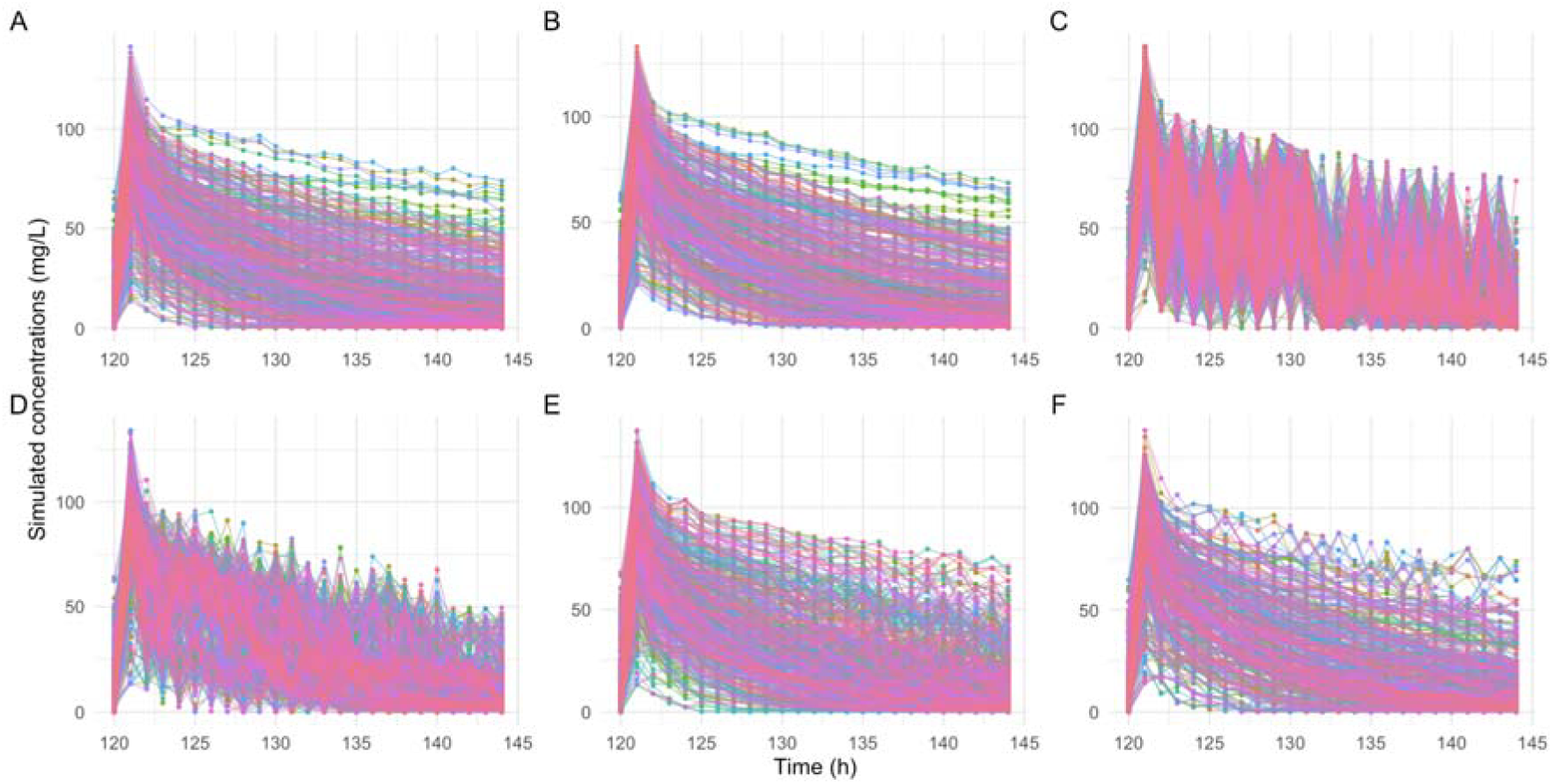
Visual predictive evaluation of longitudinal pharmacokinetic profiles. Spaghetti plots comparing the original simulated daptomycin concentration-time profiles (A) with one synthetic dataset generated by algorithms, including the modified AVATAR method (B), Conditional Tabular Generative Adversarial Networks (CTGAN) (C), Tabular Variational Auto-Encoders (TVAE) (D), Gaussian Copula (E), and Synthpop (F). Colour solid lines represent individual concentration-time trajectories. This visual assessment highlights the varying capacities of generative algorithms to reproduce the structural decay and variability inherent to the two-compartment kinetics of daptomycin.

**Figure 2:**
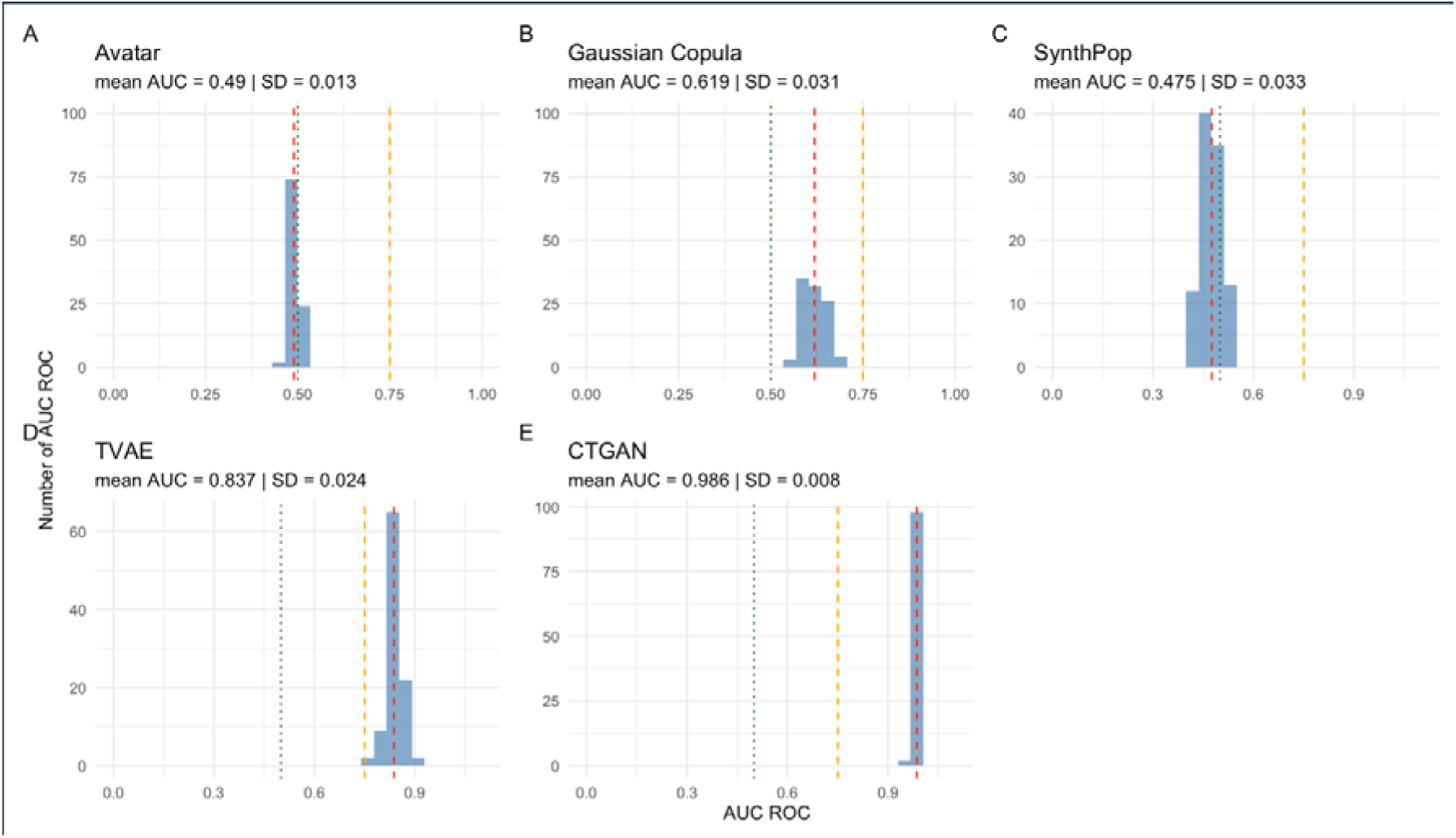
Macroscopic statistical indistinguishability screening across all generative algorithms. Histograms displaying the distribution of the Area Under the Receiver Operating Characteristic Curve (AUC ROC) values obtained from a logistic regression classifier across 100 independent synthetic datasets for each of the five evaluated algorithms: Modified AVATAR (A), Gaussian Copula (B), Synthpop (C), Tabular Variational Autoencoder (TVAE) (D), and Conditional Tabular Generative Adversarial Network (CTGAN) (E). The vertical green dotted line represents the theoretical ideal of perfect statistical indistinguishability (AUC ROC = 0.5), indicating that the classifier performs no better than random chance. The vertical red dashed line indicates the empirical mean AUC ROC across the 100 iterations for each respective method. The vertical orange dashed line represents the pre-specified eligibility threshold (AUC ROC = 0.75), above which algorithms were deemed to produce macroscopically non-representative synthetic populations and were excluded from subsequent population pharmacokinetic modelling. AVATAR (A) and Synthpop (C) demonstrated near-perfect indistinguishability (mean AUC ROC = 0.490 and 0.475, respectively), while Gaussian Copula (B) showed acceptable but partial indistinguishability (mean AUC ROC = 0.619). Conversely, TVAE (D) and CTGAN (E) exceeded the exclusion threshold (mean AUC ROC = 0.837 and 0.986, respectively), confirming macroscopic structural non-representativeness and disqualifying them from further evaluation. Abbreviations: AUC, Area Under the Curve; ROC, Receiver Operating Characteristic; TVAE, Tabular Variational Autoencoder; CTGAN, Conditional Tabular Generative Adversarial Network.

**Figure 3:**
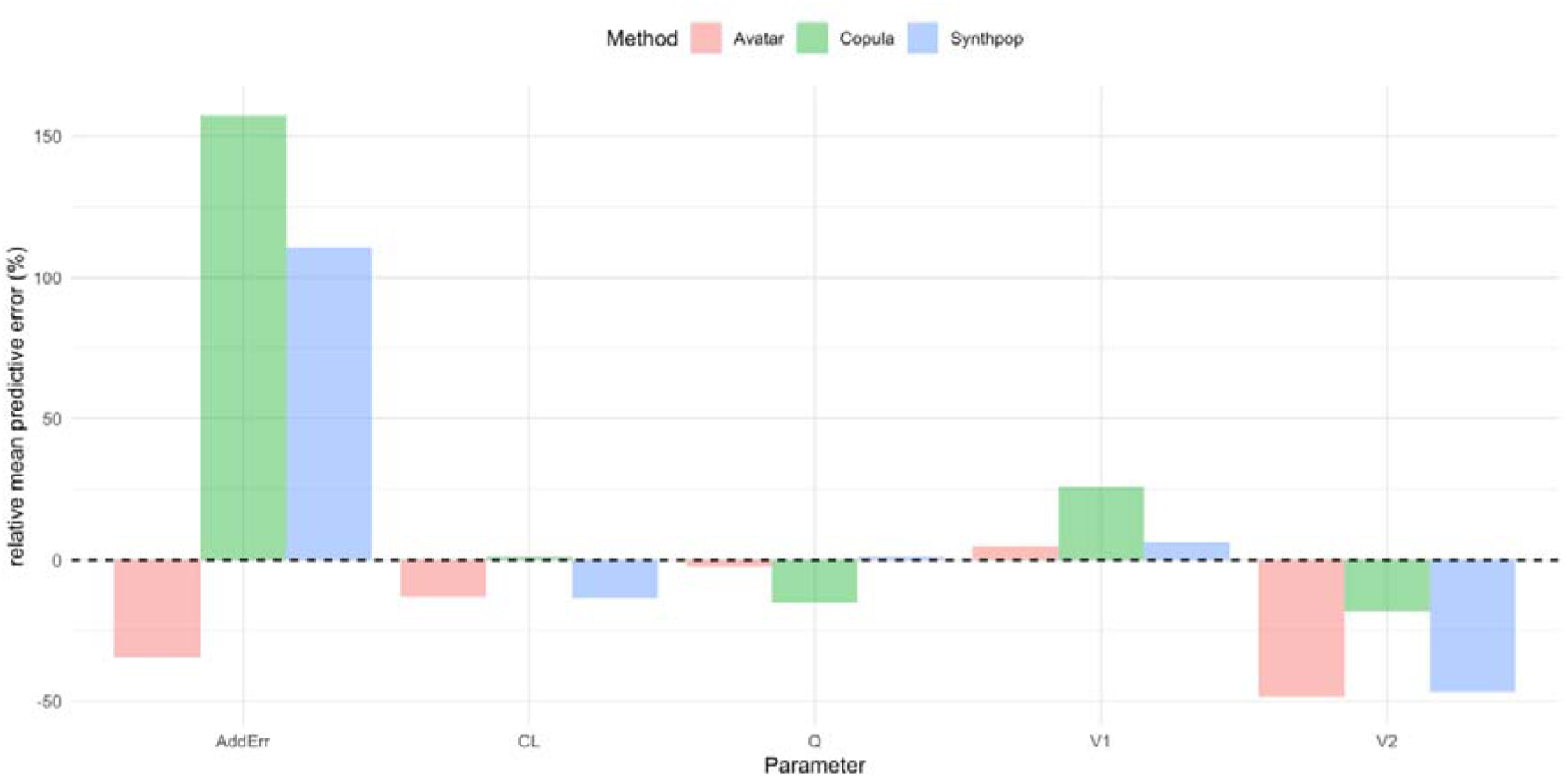
Relative bias in typical population pharmacokinetic parameters estimated from synthetic datasets. Barplot comparing the mean relative bias (%) for the structural pharmacokinetic parameters and the additive residual error model across the 100 synthetic datasets generated by AVATAR, Gaussian Copula, and Synthpop algorithms. Parameters were re-estimated using the First-Order Conditional Estimation with Interaction (FOCEi) algorithm with fixed covariate coefficients to isolate structural inference distortion. The dashed black line (y = 0) represents the reference values derived from the original simulated population. Note that the Conditional Tabular Generative Adversarial Network (CTGAN) and Tabular Variational Autoencoder (TVAE) method was excluded due to systematiic algorithmic non-convergence. Abbreviations: CL, clearance; V1, central volume of distribution; V2, peripheral volume of distribution; Q, inter-compartmental clearance; AddErr, additive residual error.

**Figure 4:**
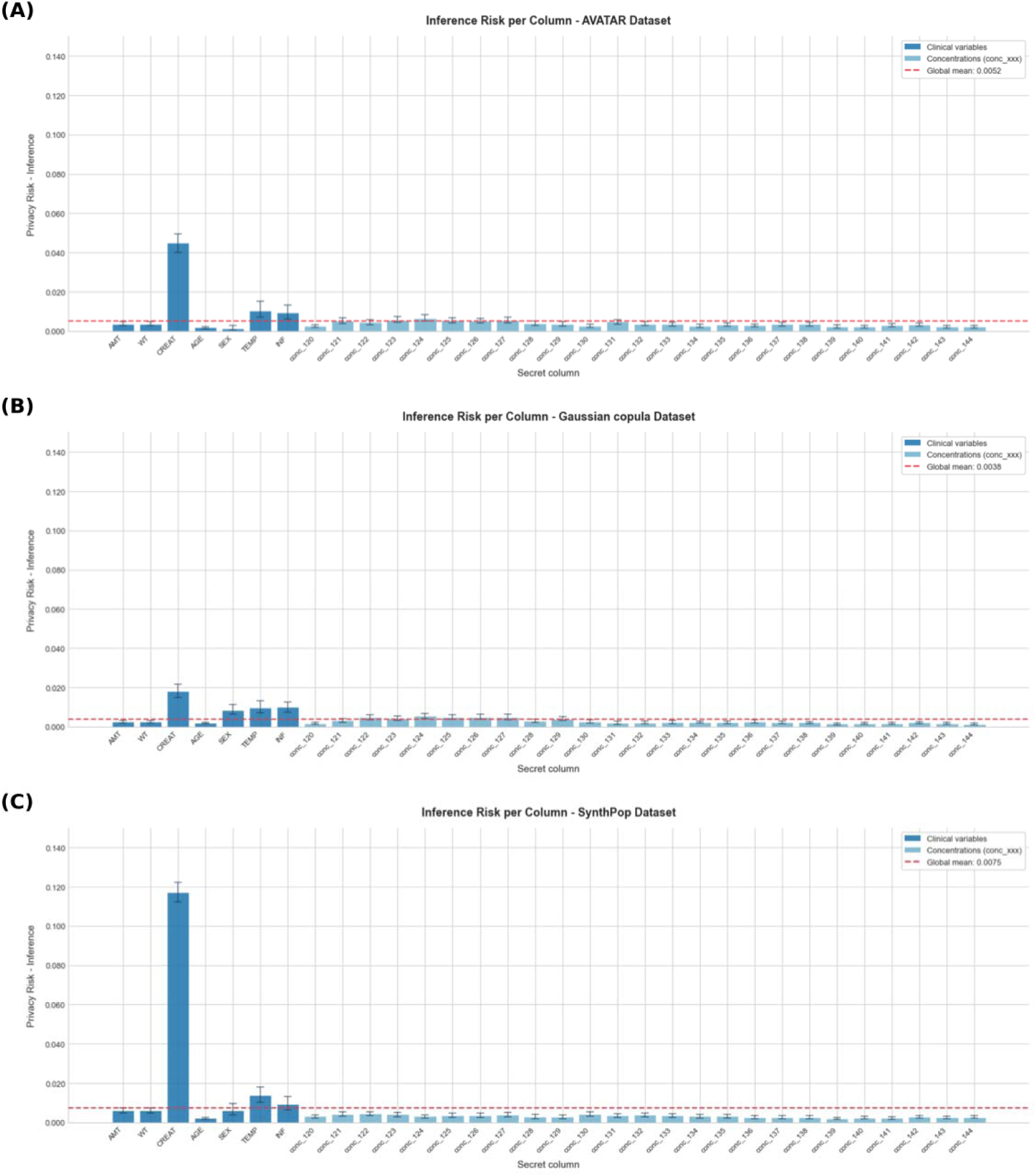
V**ariable-level inference privacy risk across convergent generative algorithms.** Bar plots illustrating the per-column inference privacy risk estimated using the Anonymeter “InferenceEvaluator” (n of attacks = 500) across the 100 independent synthetic subsets, for AVATAR (A), Gaussian Copula (B), and Synthpop (C). Error bars represent 95% bootstrap confidence intervals (n_resamples = 9,999). The dashed red line indicates the theoretical maximum risk threshold of 0.5. Across all three algorithms, creatinine (CREAT) consistently emerged as the most vulnerable variable to inference attacks, with substantially higher risk estimates compared with all other covariates and pharmacokinetic concentration variables (conc_120 to conc_144). This finding suggests that CREAT-derived variables may require additional pseudonymisation prior to synthetic data sharing, regardless of the generative algorithm employed.

**Table 1:** Demographics, clinical covariates, and pharmacokinetic characteristics of the original simulated cohort and the generative algorithm populations. Continuous variables are expressed as median [interquartile range, Q1–Q3] and categorical variables as counts (percentages). For the five synthetic methods (AVATAR, Gaussian Copula, Synthpop, TVAE, and CTGAN), demographic and clinical metrics represent the pooled distribution across the 100 independent generative iterations (resulting in a cumulative virtual population of N=50,000 per algorithm). This pooling strategy was employed to accurately capture the global generative tendency of each algorithm and isolate the absolute minimum and maximum pharmacokinetic concentration bounds. Note: Datasets generated by TVAE and CTGAN subsequently failed the population pharmacokinetic (PopPK) FOCEi re-estimation step due to extreme structural and physiological covariate distortions (e.g., biologically implausible creatinine clearance values). *Abbreviations:* CTGAN: Conditional Tabular Generative Adversarial Network; TVAE: Tabular Variational Autoencoder.

| **Generative Algorithms** |  |  |  |  |  |  |
| --- | --- | --- | --- | --- | --- | --- |
| **Characteristics** | **1. Original**<br>N = 500<sup>1</sup> | **2. AVATAR**<br>N = 50,000<sup>1</sup> | **3. Gaussian Copula**<br>N = 50,000<sup>1</sup> | **4. Synthpop**<br>N = 50,000<sup>1</sup> | **5. TVAE**<br>N = 50,000<sup>1</sup> | **6. CTGAN**<br>N = 50,000<sup>1</sup> |
| **Age (years)** | 51.2 [39.9 - 62.6] | 51.2 [42.6 - 59.9] | 51.2 [39.6 - 63.7] | 51.1 [39.7 - 62.7] | 43.2 [35.7 - 53.4] | 45.8 [32.8 - 59.2] |
| **Weight (kg)** | 81.2 [65.1 - 97.2] | 80.4 [66.3 - 95.3] | 79.9 [64.0 - 100.0] | 81.1 [65.1 - 97.0] | 83.1 [73.3 - 94.5] | 96.2 [75.0 - 118.8] |
| **Sex** |  |  |  |  |  |  |
| Female | 204 (41%) | 20,925 (42%) | 20,322 (41%) | 20,081 (40%) | 12,342 (25%) | 21,459 (43%) |
| Male | 296 (59%) | 29,075 (58%) | 29,678 (59%) | 29,919 (60%) | 37,658 (75%) | 28,541 (57%) |
| **Creatinine Clearance (mL/min)** | 140.1 [78.0 - 238.9] | 160.3 [95.1 - 273.3] | 130.7 [72.7 - 230.1] | 167.7 [89.1 - 275.6] | 156.2 [106.7 - 262.5] | 253.4 [140.1 - 446.3] |
| **Temperature (°C)** | 37.4 [36.8 - 37.9] | 37.3 [37.0 - 37.8] | 37.3 [36.8 - 38.0] | 37.4 [36.8 - 37.9] | 37.4 [36.8 - 37.8] | 37.4 [36.8 - 38.2] |
| **Infection Status** | 232 (46%) | 22,387 (45%) | 23,330 (47%) | 23,029 (46%) | 20,693 (41%) | 26,510 (53%) |
| **Patient Median Conc. (mg/L)** | 22.4 [10.5 - 39.4] | 21.6 [10.9 - 39.1] | 22.8 [10.6 - 39.5] | 23.0 [11.6 - 39.0] | 23.2 [14.2 - 36.6] | 28.4 [22.7 - 34.1] |
| **Absolute Minimum Conc. (mg/L)** | 0.0 | 0.0 | 0.0 | 0.0 | 0.0 | 0.0 |
| **Absolute Maximum Conc. (mg/L)** | 141.3 | 139.8 | 141.3 | 141.3 | 141.3 | 141.3 |

<sup>1</sup>Median [Q1 - Q3]; n (%); Min; Max

### Statistical Indistinguishability and Algorithm Pre-selection

CTGAN and TVAE exhibited high and consistent macroscopic detectability, with mean AUC ROC values of 0.986 ± 0.008 (median 0.987, IQR [0.982–0.990]) and 0.837 ± 0.024 (median 0.838, IQR [0.821–0.851]), respectively. Both algorithms exceeded the 0.75 threshold across virtually all 100 iterations, confirming that their synthetic populations were structurally non-representative of the original cohort — a finding consistent with the biological implausibility already identified at the covariate inspection stage. Both were therefore excluded from further NLME evaluation.

Synthpop, AVATAR, and Gaussian Copula all passed the macroscopic indistinguishability criterion. AVATAR demonstrated the closest approximation to statistical chance, with a mean AUC ROC of 0.490 ± 0.013 (median 0.490, IQR [0.480–0.500], range [0.461–0.516]), indicating that the adversarial classifier was unable to separate synthetic from original records beyond random performance. Synthpop showed a comparable profile, with a mean AUC ROC of 0.475 ± 0.033 (median 0.471, IQR [0.455–0.502], range [0.406–0.546]). Gaussian Copula achieved a mean AUC ROC of 0.619 ± 0.031 (median 0.616, IQR [0.594–0.645], range [0.553–0.695]), indicating acceptable but less complete macroscopic indistinguishability. All three algorithms accordingly proceeded to the subsequent NLME modelling and privacy evaluation steps.

### Population Pharmacokinetic Parameter Re-estimation and Structural Bias

To establish a rigorous comparative baseline, the original simulated dataset was first subjected to the FOCEi re-estimation process. This yielded the reference typical population parameters (0_TV_ ): CL = 0.815 L/h, V_l_ = 4.78 L, V_2_ = 3.17 L, Q = 3.51 L/h, and an additive residual error of 1.42 mg/L. The performance of the converging generative algorithms (AVATAR, Gaussian Copula, and Synthpop) was evaluated by calculating the relative bias based on these re-estimated baseline values (Table 2).

**Table 2:** Population Pharmacokinetic Parameters Estimates and Relative Bias by Generative Algorithms. *Abbreviations:* CL: clearance; V1: central volume of distribution; V2: peripheral volume of distribution; Q: inter-compartmental clearance; AddErr: additive residual error; CTGAN: Conditional Tabular Generative Adversarial Network; TVAE: Tabular Variational Autoencoder.

| Parameter | Original (Re-estimated) | AVATAR | Bias (%) | Gaussian Copula | Bias (%) | SynthPop | Bias (%) |
| --- | --- | --- | --- | --- | --- | --- | --- |
| CL (L/h) | 0.815 | 0.710(0.012) | -12.9 | 0.823(0.03) | +1.0 | 0.706(0.031) | -13.4 |
| V1 (L) | 4.78 | 5.02(0.07) | +5.0 | 6.02(0.44) | +25.9 | 5.08(0.16) | +6.3 |
| V2 (L) | 3.17 | 1.64(0.01) | -48.3 | 2.59(0.174) | -18.3 | 1.69(0.37) | -46.7 |
| Q (L/h) | 3.51 | 3.43(0.038) | -2.3 | 2.98(0.27) | -15.1 | 3.55(0.15) | +1.1 |
| AddErr (mg/L) | 1.42 | 0.933(0.133) | -34.3 | 3.65(0.31) | +157.0 | 2.99(3.11) | +110.6 |
| Mean(SD) |  |  |  |  |  |  |  |

The parameter re-estimation revealed distinct, algorithm-specific compensatory mechanisms within the nonlinear mixed-effects modelling framework. The modified AVATAR systematically reduced the intra-individual variability. This was evidenced by a -34.3% drop in the estimated additive residual error (0.93 mg/L vs. 1.42 mg/L). To compensate for this artificially smooth longitudinal trajectory, the FOCEi algorithm structurally shifted the variance, preserving the primary early-phase distribution parameters yielding excellent estimates for V_l_ (+5.0% bias) and Q (−2.3% bias), but severely deflating the V_2_ by -48.3% (1.64 L). AVATAR also slightly underestimated the systemic CL by -12.9%.

Conversely, the Gaussian Copula exhibited a diametrically opposed structural bias. While it captured the global average exposure almost perfectly, yielding an accurate CL estimate (0.823 L/h, +1.0% bias), its reliance on a global correlation matrix rather than continuous temporal logic injected important stochastic noise into the concentration-time profiles. Consequently, the FOCEi algorithm was forced to increase the additive residual error by an +157.0% (3.65 mg/L). To mathematically accommodate this noise during the initial distribution phase, the model artificially diluted the simulated drug, overestimating VLJ by +25.9% (6.02 L) and underestimating VLJ by -18.3% (2.59 L). Additionally, Q was underestimated by -15.1% (2.98 L/h).Finally, Synthpop produced a parameter profile nearly identical to modified AVATAR (CL bias of nearly 15%, V_l_ bias of +6.3% and V_2_ bias of - 46.7%). However, contrary to AVATAR, the additive residual error was increased by +110.6% (2.99 mg/L).

#### Attack-based Privacy Risk Quantification

To extend the adversarial AUC ROC findings with a granular, mechanistic assessment of re-identification risk, a complementary attack-based evaluation was performed using the Anonymeter framework across three paradigms: Singling Out, Linkability, and Inference (Table 3).

**Table 3:** Attack-based privacy risk estimates (Anonymeter framework) for the three convergent generative algorithms. Singling Out: multivariate analysis; Linkability: demographic vs. pharmacokinetic variable partition; risks expressed as mean over 100 subsets; 95% CIs obtained by non-parametric bootstrap (n_resamples = 9,999).

| Algorithm | Singling Out Risk | 95% CI | Linkability Risk | 95% CI |
| --- | --- | --- | --- | --- |
| AVATAR | 0.004 | [0.003–0.005] | 0.010 | [0.009–0.010] |
| Synthpop | 0.035 | [0.033–0.037] | 0.007 | [0.006–0.008] |
| Gaussian Copula | 0.001 | [0.001–0.002] | 0.003 | [0.003–0.004] |

Across all three convergent algorithms, absolute privacy risk estimates remained low. For Singling Out, the most stringent attack, which targets the isolation of a single individual through a unique combination of attributes, Gaussian Copula achieved the lowest risk (mean = 0.001, 95% CI [0.001– 0.002]), followed by AVATAR (0.004, [0.003–0.005]), and Synthpop (0.035, [0.033–0.037]). The markedly higher singling-out risk observed for Synthpop is consistent with its sequential CART-based generation mechanism, which may preserve rare covariate combinations and thereby render certain individuals more easily isolable in the synthetic space.

For Linkability, testing whether an adversary holding independent knowledge of demographic and pharmacokinetic variables could link them across the synthetic dataset — Gaussian Copula again demonstrated the lowest risk (0.003, [0.003–0.004]), followed by Synthpop (0.007, [0.006–0.008]) and AVATAR (0.010, [0.009–0.010]).

Variable-level inference risk analysis revealed a consistent pattern across all three convergent algorithms: creatinine (CREAT) was systematically identified as the most vulnerable variable, with mean inference risks of approximately 0.12 for Synthpop, 0.045 for Gaussian Copula, and 0.018 for AVATAR. Secondary vulnerability was observed for temperature (TEMP) and infection status (INF), while age (AGE) showed near-zero risk across all methods. Importantly, all 26 pharmacokinetic concentration variables (conc_120 to conc_144) demonstrated consistently negligible inference risk regardless of the algorithm, suggesting that the longitudinal PK structure is well-protected against attribute inference attacks.

### Clinical Impact Assessment of PopPK model developed based on synthetic data

Forward deterministic simulations were performed for three distinct virtual patient profiles to translate the structural parameter biases identified above into clinically meaningful pharmacokinetic consequences in the context of MIPD. Results are presented in Table 4 and Figure 5. Synthpop results are additionally reported for illustrative purposes to characterise the clinical consequences of proceeding with a structurally plausible but privacy-compromised algorithm.

**Figure 5:**
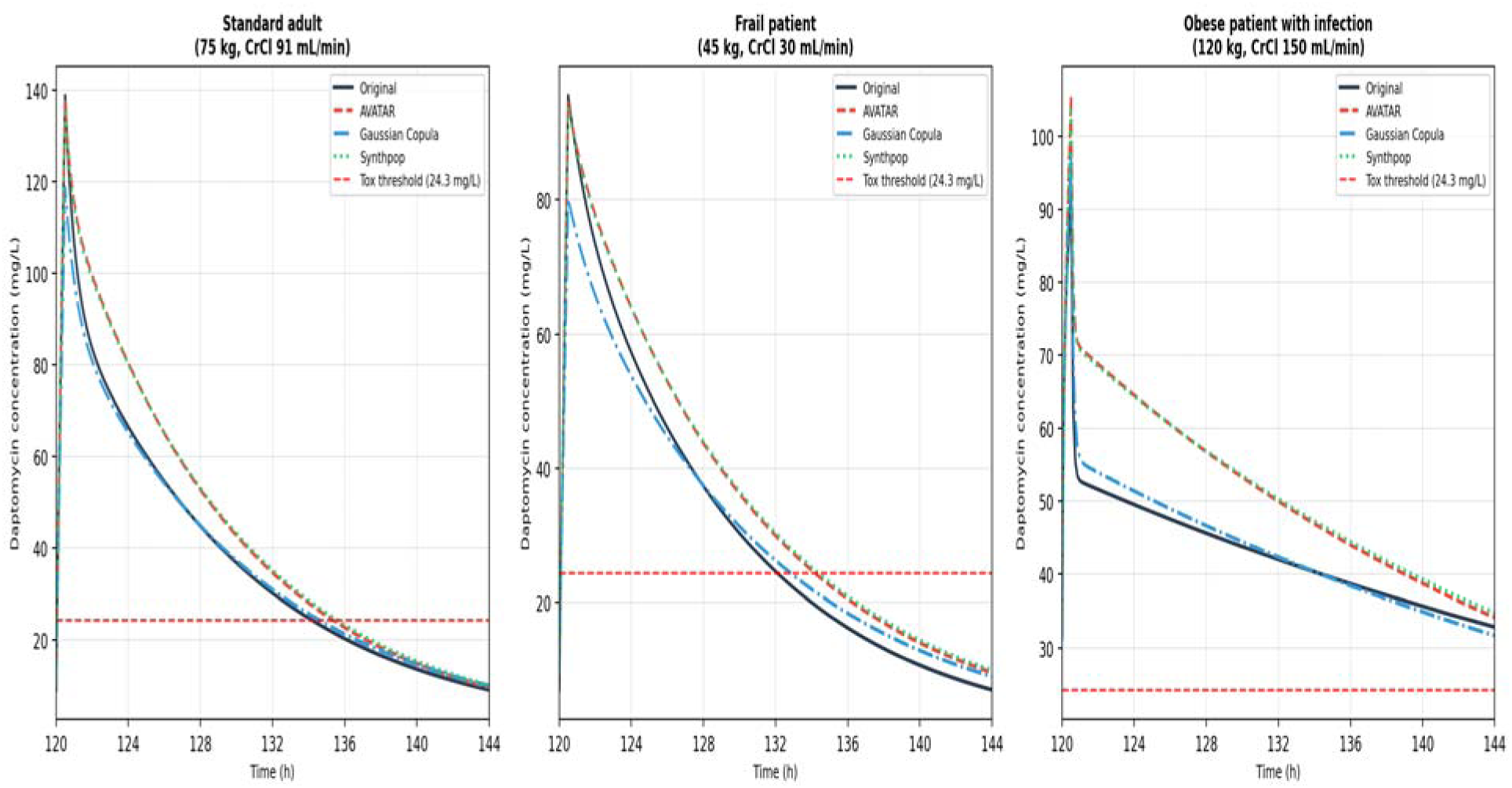
Clinical pharmacokinetic simulation of steady-state daptomycin profiles derived from synthetic-data-based population models. Steady-state concentration-time profiles (120–144 hours) simulated for three distinct virtual patient profiles under a standard daptomycin regimen of 10 mg/kg every 24 hours administered as a 30-minute intravenous infusion. Profiles were generated using typical population pharmacokinetic parameters (ETA = 0) re-estimated from the original simulated dataset (Original, black solid line), and from the three convergent generative algorithms: Modified AVATAR (red dashed line), Gaussian Copula (blue dash-dot line), and Synthpop (green dotted line). Patient profiles: (A) standard adult (75 kg, CrCl 91 mL/min, no infection); (B) frail low-weight patient (45 kg, CrCl 30 mL/min, no infection); (C) obese patient with systemic infection (120 kg, CrCl 150 mL/min, INF = 1). The horizontal red dashed line indicates the established daptomycin skeletal muscle toxicity threshold (Cmin > 24.3 mg/L). Gaussian Copula systematically underestimated peak concentrations across profiles (A) and (B), while AVATAR- and Synthpop-derived models overestimated both Cmax and Cmin in profile (C), compounding the accumulation risk inherent to this patient. *Abbreviations:* CrCl, creatinine clearance; INF, systemic infection status; Cmax, maximum steady-state concentration; Cmin, minimum steady-state concentration.

**Table 4:** Steady-state Cmax and Cmin predictions by patient profile and generative model (10 mg/kg q24h, 30-min IV infusion, simulated at 120–144 hours).

| Patient Profile | Model | C<sub>max</sub> (mg/L) | C<sub>min</sub> (mg/L) | Toxicity risk |
| --- | --- | --- | --- | --- |
| **Standard adult**<br>(75 kg, CrCl 91 mL/min) | Original | 138.85 | 9.24 | — |
|  | AVATAR | 137.56 | 9.80 | — |
|  | Gaussian Copula | 118.77 | 10.13 | — |
|  | Synthpop | 136.23 | 10.23 | — |
| **Frail patient**<br>(45 kg, CrCl 30 mL/min) | Original | 95.53 | 7.04 | — |
|  | AVATAR | 94.55 | 9.58 | — |
|  | Gaussian Copula | 80.23 | 9.01 | — |
|  | Synthpop | 93.86 | 9.92 | — |
| **Obese with infection**<br>(120 kg, CrCl 150 mL/min, INF = Yes) | Original | 92.01 | 31.69 | Above threshold (>24.3 mg/L) |
|  | AVATAR | 105.51 | 33.73 | Above threshold (>24.3 mg/L) |
|  | Gaussian Copula | 96.93 | 30.89 | Above threshold (>24.3 mg/L) |
|  | Synthpop | 104.17 | 34.29 | Above threshold (>24.3 mg/L) |

C<sub>min</sub> toxicity threshold: > 24.3 mg/L. All simulations performed with ETA = 0 (typical patient, no between-subject variability).

For the standard adult patient (75 kg, CrCl 91 mL/min), the original re-estimated model predicted a steady-state Cmax of 138.85 mg/L and a Cmin of 9.24 mg/L, remaining well below the toxicity threshold (Cmin > 24.3 mg/L). AVATAR and Synthpop reproduced the Cmax with good accuracy (137.56 mg/L, -0.9% and 136.23 mg/L, -1.9%, respectively), with slightly elevated but clinically acceptable Cmin values (9.80 and 10.23 mg/L). In contrast, the Gaussian Copula model substantially underestimated Cmax (118.77 mg/L, -14.5% relative to the original), a direct consequence of VLJ overestimation inflating the apparent distribution volume and blunting the peak concentration.

For the frail patient (45 kg, CrCl 30 mL/min), the original model predicted a Cmax of 95.53 mg/L and a Cmin of 7.04 mg/L. The Gaussian Copula again markedly underestimated Cmax (80.23 mg/L, - 16.0%), a clinically relevant deviation that could prompt unnecessary empirical dose escalation in a patient already at elevated risk of drug accumulation. AVATAR and Synthpop reproduced Cmax with reasonable accuracy (94.55 and 93.86 mg/L) but predicted higher Cmin values (9.58 and 9.92 mg/L versus 7.04 mg/L for the original), reflecting the reduced VLJ and slower apparent elimination driven by CL underestimation. Importantly, the toxicity threshold (Cmin > 24.3 mg/L) was not exceeded by any model in this patient profile.

For the obese patient with systemic infection (120 kg, CrCl 150 mL/min, INF = 1), the multiplicative effect of infection on VLJ (×1.93) dramatically expanded the peripheral compartment, resulting in markedly elevated Cmin values across all models. The original model predicted a Cmax of 92.01 mg/L and a Cmin of 31.69 mg/L, already exceeding the toxicity threshold. The Gaussian Copula model performed closest to the original for this profile, predicting a Cmax of 96.93 mg/L and Cmin of 30.89 mg/L. Conversely, AVATAR and Synthpop, whose deflated VLJ estimates were partially compensated by the infection multiplier, predicted a substantially higher Cmax (105.51 and 104.17 mg/L, +14.7% and +13.2% respectively) and further elevated Cmin values (33.73 and 34.29 mg/L), compounding the toxicity risk already present in this vulnerable profile. Across all four models, the Cmin exceeded 24.3 mg/L for this patient, confirming that the 10 mg/kg q24h regimen would be inappropriate regardless of the model used, but the degree of overestimation introduced by AVATAR- and Synthpop-derived models would further mislead the clinical assessment of the accumulation risk.

## Discussion

The sharing of individual patient data is essential for progress in pharmacometrics, as it enables external validation, methodological development, and the construction of more robust and generalizable PopPK models. However, IPD sharing remains strongly limited by privacy regulations, particularly the European GDPR. In this context, synthetic data generation may offer a privacy-preserving alternative, while also supporting open science by facilitating transparency, reproducibility, secondary analyses, benchmarking, and educational use without directly exposing sensitive patient-level data.

Using daptomycin as a case study, this benchmark showed that synthetic data have substantial potential for longitudinal PopPK data sharing, but that their performance depends strongly on the generative algorithm. Among the evaluated methods, only the modified AVATAR algorithm and Gaussian Copula satisfied both macroscopic indistinguishability and attack-based privacy criteria, although with distinct privacy–utility profiles.

Deep learning-based approaches, namely CTGAN and TVAE, were less suitable in this setting. FOCEi re-estimation failed to converge for the datasets generated by these methods, suggesting that the synthetic profiles were not sufficiently compatible with standard compartmental pharmacokinetic modelling. While this does not identify a single failure mechanism, it is consistent with the known data requirements of neural-network-based generators, which often require large training sets to learn stable mixed-type and longitudinal dependency structures[15]. In the present study, the training cohort of 500 patients may have been insufficient for CTGAN and TVAE to adequately reproduce concentration–time trajectories and physiological constraints.

By contrast, Gaussian Copula, Synthpop, and AVATAR showed greater robustness in this small-sample PopPK setting. Their reliance on explicit statistical dependence modelling or local/sequential data-construction strategies may explain their better ability to generate structurally plausible datasets suitable for downstream pharmacometrics analysis.

This study proposes a structured two-stage framework for evaluating synthetic data generation algorithms in pharmacometrics. First, macroscopic indistinguishability, assessed using logistic regression, serves as an eligibility filter to exclude algorithms that fail to reproduce the global multivariate structure of the source dataset. In this study, CTGAN and TVAE were excluded at this stage, whereas AVATAR, Gaussian Copula, and Synthpop showed acceptable population-level similarity.

However, global similarity was not sufficient to ensure privacy. Synthpop achieved an AUC ROC close to chance, comparable to AVATAR, yet displayed a substantially higher singling-out risk in the Anonymeter assessment. This discrepancy shows that population-level indistinguishability and individual-level privacy risks are distinct properties. A synthetic dataset may accurately reproduce the overall distribution of the original data while still preserving rare combinations of attributes that make specific individuals vulnerable to re-identification.

The attack-based privacy assessment provided a necessary complementary layer of evidence that the macroscopic indistinguishability screen alone cannot capture. Among the algorithms passing the initial screening, Gaussian Copula showed the lowest risks for both singling-out (0.001, 95% CI [0.001–0.002]) and linkability (0.003, [0.003–0.004]), likely reflecting the smoothing effect of its parametric covariance structure. AVATAR showed slightly higher but still acceptable risks (singling-out = 0.004; linkability = 0.010), whereas Synthpop’s elevated singling-out risk (0.035, [0.033–0.037]) supported its exclusion from privacy-preserving data sharing despite its satisfactory AUC ROC. Notably, although Gaussian Copula achieved the lowest absolute re-identification risk across both attack paradigms, it exhibited substantially weaker macroscopic indistinguishability (AUC ROC = 0.619) compared with AVATAR (AUC ROC = 0.490). This apparent discrepancy underlines that population-level distributional overlap and individual-level re-identification risk are orthogonal properties that cannot be inferred from one another, a synthetic dataset can be globally more detectable at the population level while simultaneously being harder to attack at the individual level. These findings emphasize that no single metric can adequately characterise the full privacy landscape of a synthetic pharmacometrics dataset, and that a multi-attack evaluation framework is required for regulatory-grade assessment. Finally, the variable-level inference analysis identified creatinine clearance as the main vulnerability across algorithms. This is pharmacologically plausible, as renal function is a continuous, information-rich covariate that directly contributes to pharmacokinetic variability in the daptomycin model. Its consistent identification across methods suggests that this risk is primarily driven by the structure of the clinical pharmacokinetic dataset rather than by a specific generative algorithm. Additional protection strategies, such as targeted generalization or pseudonymisation of renal function variables, should therefore be considered before releasing synthetic pharmacometrics data.

While the primary exposure parameters, such as CL and V_l_ were estimated with reasonable accuracy by the two eligible algorithms, the estimation of the V_2_ and the residual error model revealed significant and algorithm-specific compensatory mechanisms. In the context of nonlinear mixed-effects modelling, algorithms tend to compensate for structural variance distortions by adjusting unexplained variability. The Gaussian Copula, while capturing multivariate dependencies, introduces a stochastic noise that disrupts the longitudinal signal; the FOCEi algorithm absorbs this noise by significantly inflating the additive residual error (+157.0%) and overestimating V1(+25.9%). Conversely, the AVATAR method, which calculates a stochastic barycentre of nearest neighbours in a PCA space, acts as a smoothing filter. This smoothing technique has been shown to preserve the global structural trajectory while concomitantly reducing overall variance, resulting in a significant underestimation of V2 (−48.3%) and a deflated residual error.

These algorithm-specific structural biases resulted in clinically distinct pharmacokinetic prediction errors across the simulated patient profiles. In standard and frail patients, Gaussian Copula-derived models consistently underestimated Cmax, mainly because overestimation of VLJ increased the apparent central distribution volume. In a MIPD setting, such underestimation could lead to unnecessary dose escalation, with a potential risk of accumulation and toxicity, particularly in patients with impaired renal function. By contrast, AVATAR- and Synthpop-derived models reproduced Cmax more accurately in these profiles but tended to overpredict Cmin, consistent with CL underestimation and marked VLJ deflation. This could falsely suggest drug accumulation and potentially prompt unwarranted dose reduction or interval extension.

The obese patient with systemic infection represented the most challenging scenario. In this profile, the infection-related increase in VLJ markedly expanded peripheral distribution and resulted in Cmin values above the toxicity threshold across all models, including the original one. This confirms that the 10 mg/kg q24h regimen would be inappropriate for this patient profile regardless of the model used, highlighting the need for individualized dose adjustment. However, AVATAR- and Synthpop-derived models further amplified the predicted exposure, overestimating both Cmax and Cmin, which could exaggerate the perceived toxicity risk. In contrast, Gaussian Copula provided the closest predictions to the original model in this profile, likely because its preserved CL estimate partially compensated for inaccuracies in distribution parameters.

Overall, these simulations show that the clinical impact of synthetic-data-derived model bias is patient-dependent and algorithm-specific. Gaussian Copula may carry a greater risk of underestimation-driven dose escalation in standard and frail patients, whereas AVATAR and Synthpop may overestimate accumulation in patients with complex distribution changes. Neither direction of error is clinically neutral for daptomycin, given the importance of AUC for efficacy and the association between elevated Cmin and toxicity.

These findings should nevertheless be interpreted in the context of real-world MIPD workflows. In clinical practice, PopPK models are used as Bayesian priors and are subsequently updated using observed therapeutic drug monitoring data. MAP estimation and empirical Bayes estimates can therefore partially correct initial structural bias once patient-specific concentrations become available. Synthetic-data-derived models may thus affect initial dose recommendations, but subsequent TDM-guided adaptation is expected to reduce part of the associated clinical risk.

A major strength of this study is the rigor of its evaluation framework, based on 100 independent iterations. This design allowed us to quantify intra-algorithm variability, an aspect that is often insufficiently addressed in synthetic data studies. In addition, the preservation of the covariate correlation structure supported the decision to fix covariate effects during FOCEi re-estimation, thereby focusing the analysis on the impact of each generative algorithm on structural pharmacokinetic parameters while limiting confounding from unstable covariate estimation.

Several limitations should nevertheless be acknowledged. First, this study relied on a rich and perfectly aligned steady-state sampling design, whereas real-world TDM datasets are typically sparse, unbalanced, and collected at heterogeneous sampling times. Under such conditions, algorithms requiring a strict wide-format structure, such as AVATAR, may be less directly applicable unless substantial preprocessing or imputation is performed. Future work should therefore evaluate these methods in sparse and irregular PopPK datasets that better reflect routine clinical practice.

This study also shows that synthetic data generation is not a neutral transformation of the original dataset. Each algorithm introduced specific structural biases, with direct implications for downstream pharmacometrics inference. Within the sample size range typical of clinical PopPK studies, deep learning-based methods such as TVAE and CTGAN showed limited ability to capture rigid physiological and longitudinal constraints, leading frequently to model non-convergence. Their current out-of-the-box applicability to small tabular PopPK datasets therefore appears limited, although they may remain relevant in larger datasets or with architectures specifically designed for longitudinal pharmacokinetic data.

Among the convergent methods, a clear privacy–utility trade-off emerged. The modified AVATAR algorithm showed the most favourable privacy profile and appeared promising for secure multicentre data sharing, but its neighbourhood-based generation process tended to smooth longitudinal profiles, leading to underestimation of peripheral volume and residual variability. Gaussian Copula better preserved some global pharmacokinetic features, particularly exposure-related parameters, but introduced greater stochastic noise and offered weaker privacy protection. Synthpop generated structurally plausible data and passed the macroscopic indistinguishability screen, but its elevated singling-out risk made it unsuitable for privacy-preserving data sharing in this setting.

This latter finding underlines a key methodological point: population-level similarity is not sufficient to establish privacy. Synthpop’s sequential CART-based generation mechanism appeared able to reproduce marginal and joint distributions at the population level, explaining its satisfactory AUC ROC, while still preserving local covariate structures that exposed individuals to targeted singling-out attacks. This reinforces the need to combine utility metrics with explicit attack-based privacy assessments when evaluating synthetic datasets for pharmacometrics data sharing.

Overall, synthetic PopPK datasets may provide immediate value for collaborative research, external evaluation of models, education, and methodological benchmarking, and may also support open science by facilitating the dissemination and secondary reuse of datasets underlying published studies. However, our mechanistic evaluation suggests that caution is needed before using models derived from synthetic data directly for a priori empirical MIPD. In practice, any such use should ideally remain coupled with therapeutic drug monitoring and Bayesian forecasting, which are essential to update prior model predictions with observed patient data, mitigate generative biases, and support safe dose individualization.

Funding information: This work is part of the DIGPHAT project which was supported by a grant from the French government, managed by the National Research Agency (ANR), under the France 2030 program, reference ANR-22-PESN-0017. This work was also supported by a grant from the ARS Nouvelle Aquitaine Regional APMT project.

### Use of artificial intelligence tools

Artificial intelligence tools were used to support manuscript preparation. Claude was used for language editing, text formulation, and improvement of English scientific style. All scientific content, interpretations, references, and final wording were critically reviewed and validated by the authors, who remain fully responsible for the manuscript.

## Data Availability

All data produced in the present study are available upon reasonable request to the authors.

